# Estimating the total size of coronavirus epidemic in Algeria via different approaches

**DOI:** 10.1101/2020.07.29.20164509

**Authors:** Mohamed Rezki

## Abstract

In this paper, several techniques and models proposed the spread of coronavirus (Covid-19) and determines approximately the final number of coronavirus infected cases as well as infection point (peak time) in Algeria. To see the goodness of the predicting techniques, a comparative study was done by calculating error indicators such as Root-Mean-Square Error (RMSE) and the sum of squared estimate of errors (SSE). The main technique used in this study is the logistic growth regression model widely used in epidemiology. The results only relate to the two months from the beginning of the epidemic in Algeria, which should be readjusted by integrating the new data over time, because hazardous parameters like possible relaxations (decrease of vigilance or laxity of society) can affect these results and generally cause a time lag in the curve. Hence, a re-estimation of the curves is always requested.

## 1. INTRODUCTION

It has become very urgent whether it was for the population or for the authorities to know the outcome of the actual pandemic (Covid-19). There remains only the pace of its evolution and the estimated total number of infectious cases that can be taken into account. A mathematical tool dedicated to epidemiology, helps public health practitioners in predicting the number of cases and deaths in the short-term, in order to plan resources and to evaluate the effectiveness of intervention strategies [1].

An epidemiological model is very useful but is always approximate because it does not depend only on the disease but it also depends on the behavior of the population, the size of the population and the different interventions made by the influential authorities.

In order to model the spread of the coronavirus epidemic, a typical method was followed as schematically represented in figure 1. This diagram is based on the fact that modelling infectious diseases and their spread in the population and the mathematical modelling of any biological system has the same principle [2].

**Figure 1.**
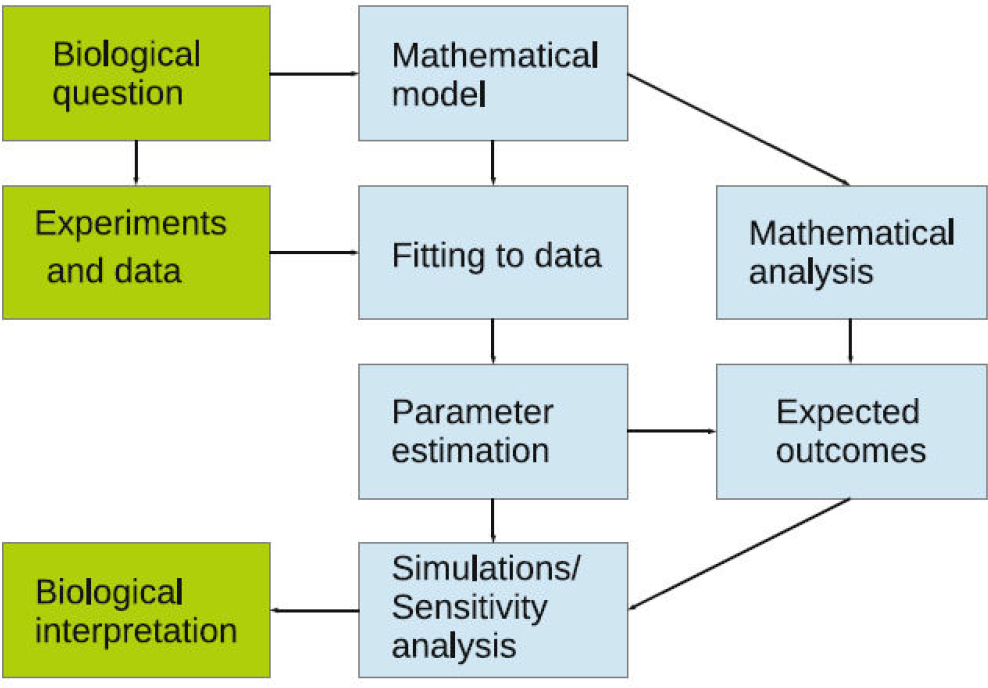
Modelling diagram [2].

From this diagram (figure1), the importance of fitting the actual data, estimation of parameters and simulation/analysis of the mathematical chosen model can be seen.

Therefore, studying the spread of coronavirus in Algeria, this work goes from a basic approach that consists on fitting the data and doing extrapolations after we follow up with a predictive study according to the logistic -regression model and finally, we end with the SIR (susceptible-infected-recovered) model.

Thus, the continuation of this paper will be as follows: Section 2 introduces the methods. Section 3 presents the different results with discussion and Section 4 gives conclusions.

## 2. METHODS

The ideal technique for predicting the output of a phenomenon is to create an equation from the data acquired. Hence, the importance of the curve fitting (linear and non-linear), compartmental models, and other methods, that can give a sense to the real data.

### 2.1 Curve fitting

The curve fitting captures the trend in the data by assigning a single function across the entire range [3]. This fitting can be linear with an equation that illustrates a straight line, its equation is f(x) = ax+b. With: a and b been the fitting parameters.

The fitting can be non-linear used for the non-linear data, such as polynomial and Gaussian with a power model.

As an example the polynomial fitting with a general order j, its equation is: f(x)=a_0_ +a_1_x +a_2_x^2^+a_3_x^3^+…..a_j_x^j^. It can be seen that the linear fitting is just a polynomial fitting with order 1.

The mean problem of this type of fitting is the possibility of having either an under-fitting (low degree of fitting) or an over-fitting (more than necessary we added noises to the data) [4].

### 2.2 Logistic model

Most epidemics grow approximately exponentially during the initial phase of an epidemic. The logistic model is a model that adds a term of saturation to the initial exponential growth [5]. This model is named model of population growth and it’s characterized in general by the following formula, called logistic formula [6]:

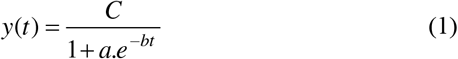

In which:

- *y(t)* is the number of cases at any given *time t*
- *C* is the limiting value, the maximum capacity for *y* (the final epidemic size, named also carrying capacity).
- *b* is the growth rate.

The maximum growth rate is at: t = *ln(a)* /b and y(t) = *C / 2*.This time is considered the peak time.

In general, if we compare this model with the curve fitting, we can observe that the logistic model is just a specific function of fitting (specific equation).

### 2.3 SIR model

In this model, we have three classes (compartments) changing with time (t): susceptible individuals (S), infected individuals (I) and removed/recovered individuals (R). The total population size *N* is the sum of the sizes of these three classes [7]:

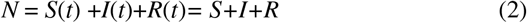

We also have:

**s(t) = S(t)/N**, the susceptible fraction of the population,

**i(t) = I(t)/N**, the infected fraction of the population,

and **r(t) = R(t)/N**, the recovered fraction of the population.

The global model which is a compartmental model can be expressed as a system of differential equations, as follows [8]:

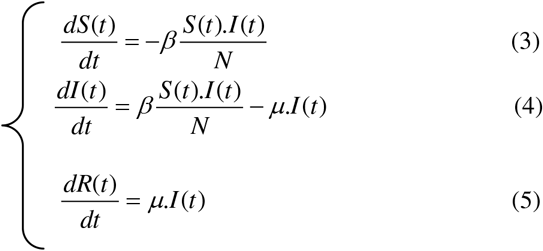

Where β is the rate of infection and μ is the rate of recovery.

This system is a system of ordinary differential equations that has analytical and numeric solutions.

## 3. RESULTS AND DISCUSSION

### 3.1 Curve fitting

The results of the curve fitting are shown in figures 2 & 3. This work is performed on data given by [9].

**Figure 2.**
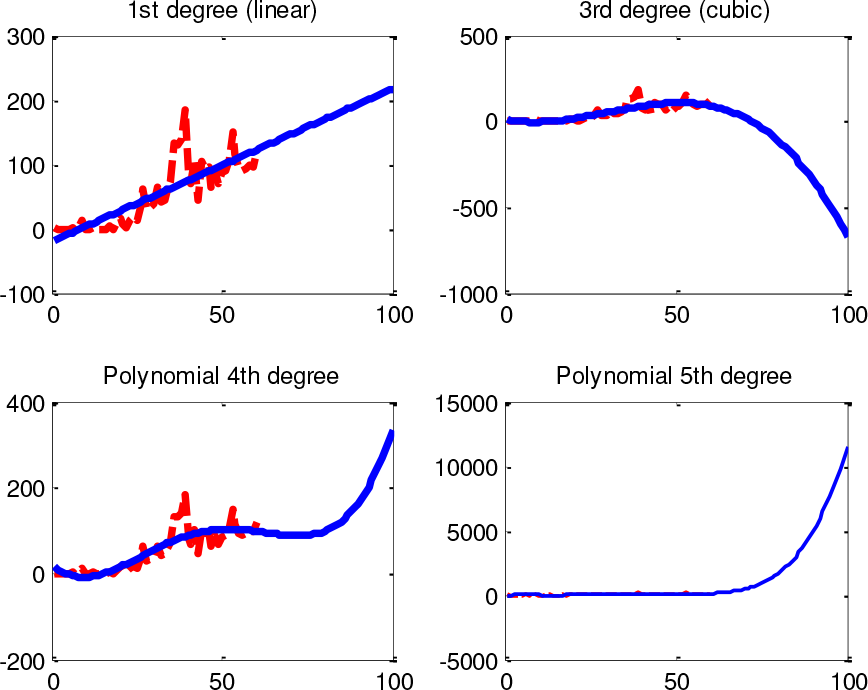
Evolution and prediction by polynomial fit of infected daily cases vs date (since 1^st^ apparition of covid on 25^th^ February)

**Figure 3.**
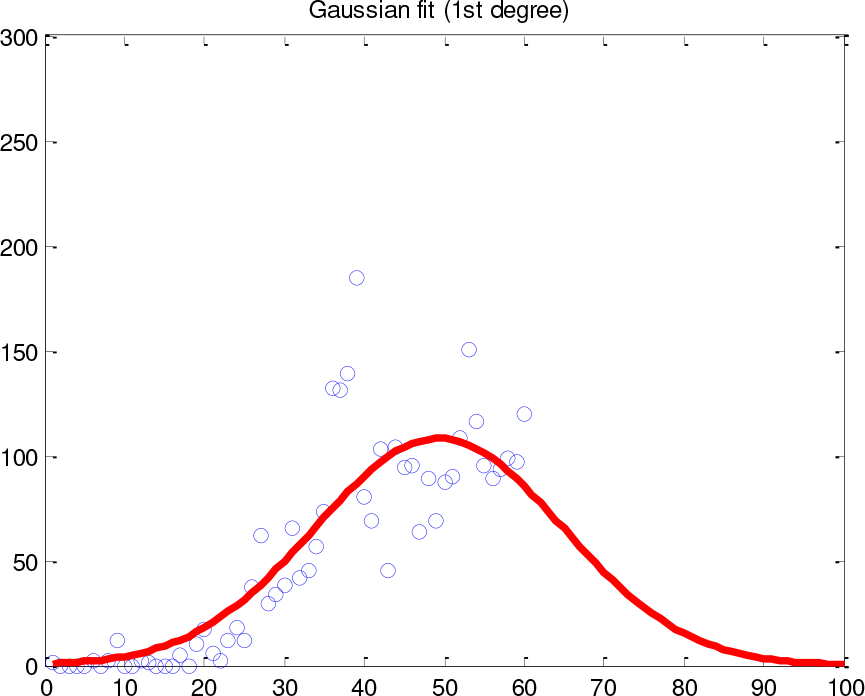
Evolution and prediction by Gaussian fit of infected daily cases vs date (since 1^st^ apparition of covid on 25^th^ February).

**Figure 4.**
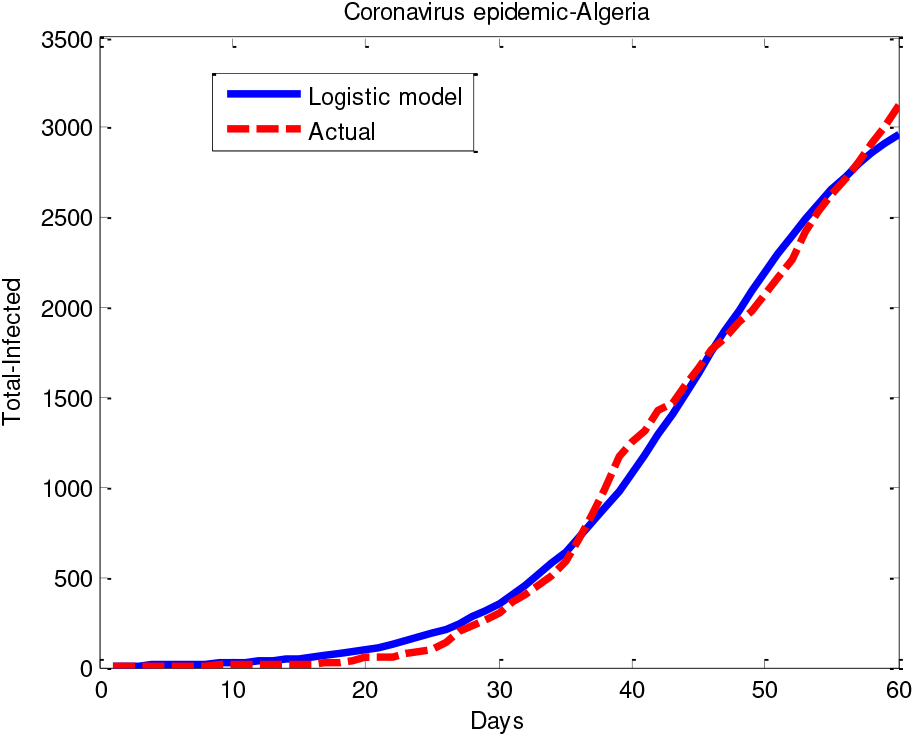
Fitting according to the logistic model (data until 25 Apr, 2020).

For the first analysis of this model and considering the shape of the epidemic which resembles a bell, the third degree of fitting (most likely for a controlled epidemic) and the fourth degree (which most resembles a hardly controlled epidemic) seem the most acceptable but only statistical calculations (of errors) can prove this. Another remark is that the cubic regression gives us an extinction of the epidemic in Algeria after 72 days (and there are no cases listed negative).

If we take a Gaussian fit (1^st^ degree) with its function as: f(x) = a1.exp (-((x-b1)/c1)^2), by execution of Matlab software, we found for our data: : a1 = 108.1, b1 = 49.35, c1 = 21.93.

It has been seen that the peak time for the Gaussian and cubic fits is around the 50^th^ day which corresponds to 14^th^ April.

Table 1 gives an error estimation of the different curve fitting types used.

**Table 1.**
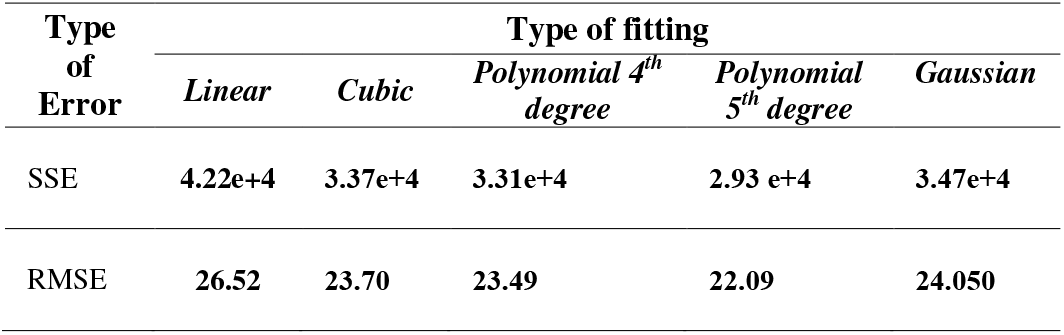
Error indicators

| Type of Error | Type of fitting |  |  |  |  |
| --- | --- | --- | --- | --- | --- |
|  | Linear | Cubic | Polynomial 4<sup>th</sup> degree | Polynomial 5<sup>th</sup> degree | Gaussian |
| SSE | 4.22e+4 | 3.37e+4 | 3.31e+4 | 2.93 e+4 | 3.47e+4 |
| RMSE | 26.52 | 23.70 | 23.49 | 22.09 | 24.050 |

The curve fitting gives just a primary estimation of the outbreak of the epidemic due to high probability of the existence of overfitting.

### 3.2 Logistic model

By applying data given by [9] the model is based on a tutorial by David Arnold [10] and implemented by Varuna De Silva [11], we get:

The results of the given model are as follows: SSE=304188.3129, RMSE=71.206, the Estimated infection point is the 31^st^ day (corresponds to 26^th^ March). The estimated carrying capacity (total size of the infected population) is about 3356.

Let’s recall that until 25^th^ April 2020 (i.e. after two months after the apparition of the coronavirus in Algeria), the situation was as expressed in figure 5.

**Figure 5.**
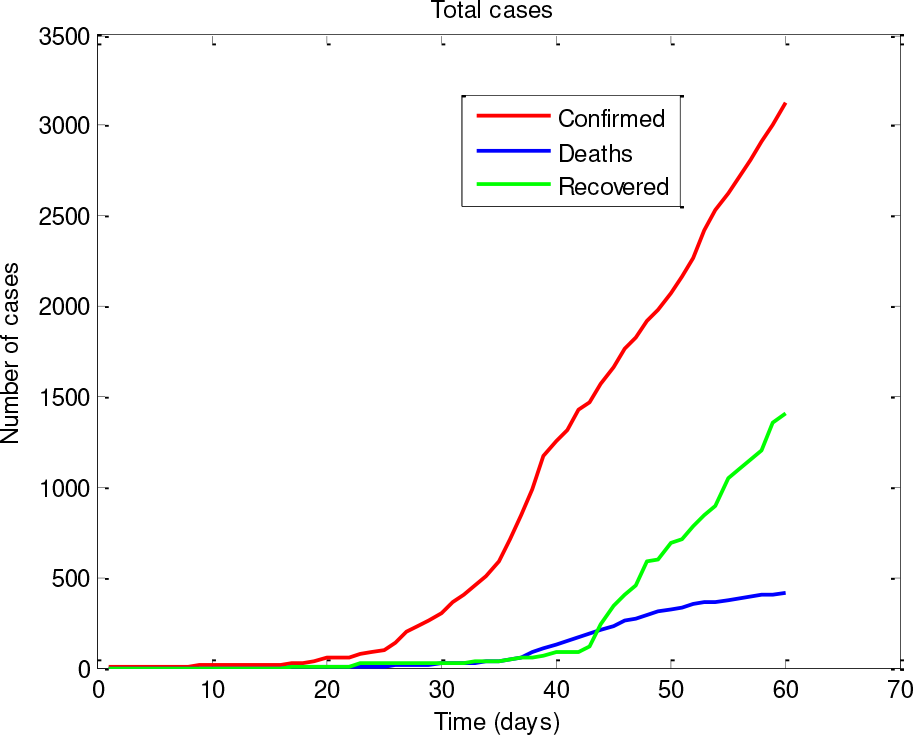
Evolution of cumulative cases-Covid-19 in Algeria (data until 25 Apr, 2020).

However, there is another classic method while exploiting the logistic model which we used in order to estimate the point of infection more precisely (ie the peak time of the epidemic), it’s the calculation of the growth factor. As we know, the growth factor at this point is equal to 1 [12] [13]. The principle can be well understood by following the graph below (figure 6 ‘a’ and ‘b’).

**Figure 6.**
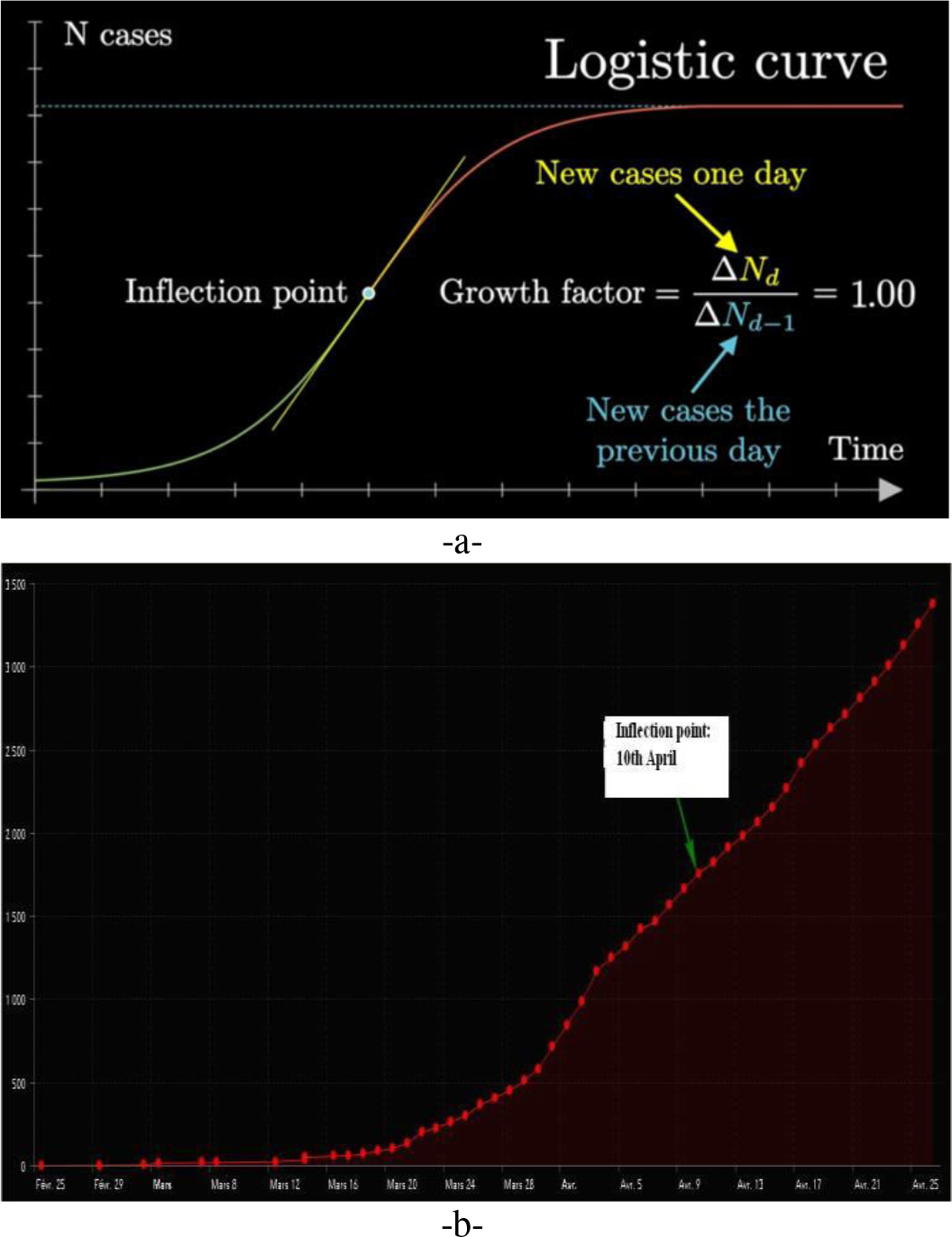
Growth factor and Covid-19 in Logistic curve: -a-Principle [12], -b-Data of Algeria [9].

By calculating the growth factor throughout the range of data acquired in order to detect the unit value, we found two values close to unity (a negative and a positive change), which are:

0.9924 (at 37th day which corresponds to 1^st^ April) and the positive change 1.0106 corresponding to the 46^th^ day (10^th^ April) from the beginning of the epidemic in Algeria. This logically gives the peak time in Algeria from 25th February to 25th April to be 10 April. Knowing, that the peak corresponds to half of carrying capacity (see the section of methods), we can easily estimate the final size of the infected population in Algeria which is: 2 × number of confirmed cases (by 10^th^ April) = 2x 1761=3522.

This result is acceptable compared to other techniques because it uses real data without prediction.

### 3.3 SIR model

In order to do a strong comparison between the methods cited above and the SIR model, we used the results of applied SIR model given by [14] and inspired from the open-source codes from Milan Batista [15] and using the daily updated COVID-19 data from Our World in Data [16].

The results of the applied SIR model are as follows:

It can be seen from figure 7 that the turning date (peak time) in Algeria is 10th April. This result confirms that from the logistics model, the estimated extinction time of the epidemic in Algeria from this study is around 8th June 2020.

**Figure 7.**
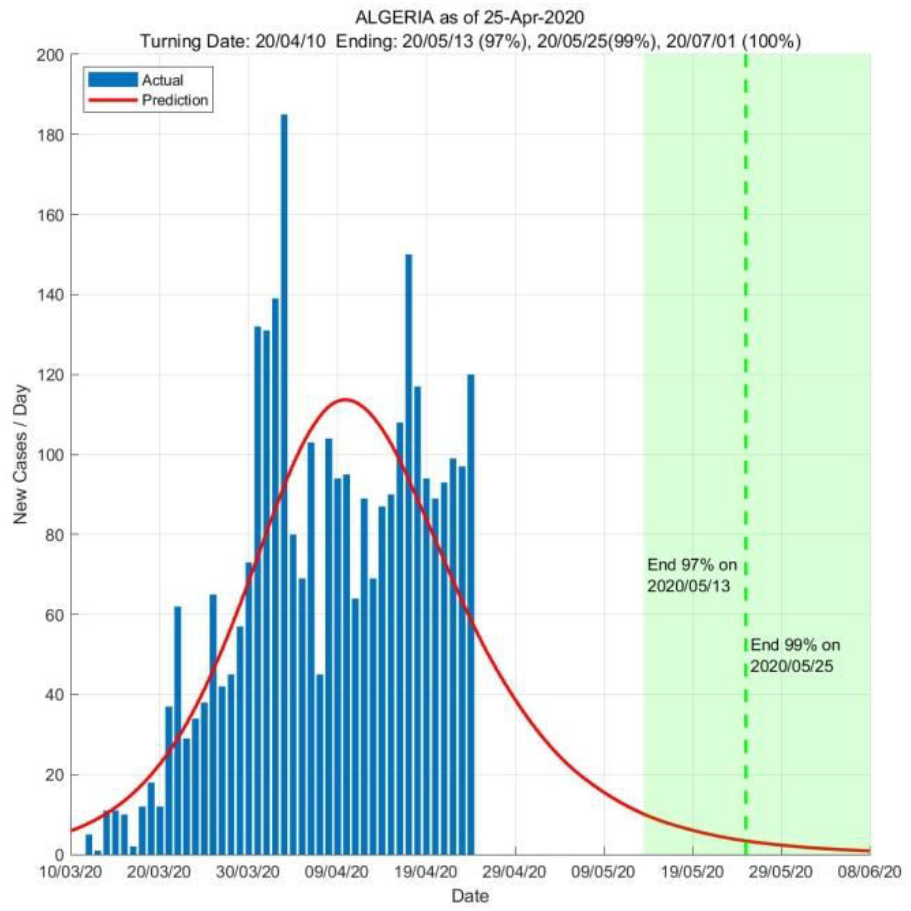
Model-Based Data-Driven Estimation of COVID-19 Life Cycle, Turning and Ending Dates in Algeria based on data as of 25 April 2020 [14].

The results can be confirmed with the study done by the ETH Zürich & EPFL, the University of Geneva exposed in figure 8 [17]. Note that the principle of this study is not based on the SIR model.

**Figure 8.**
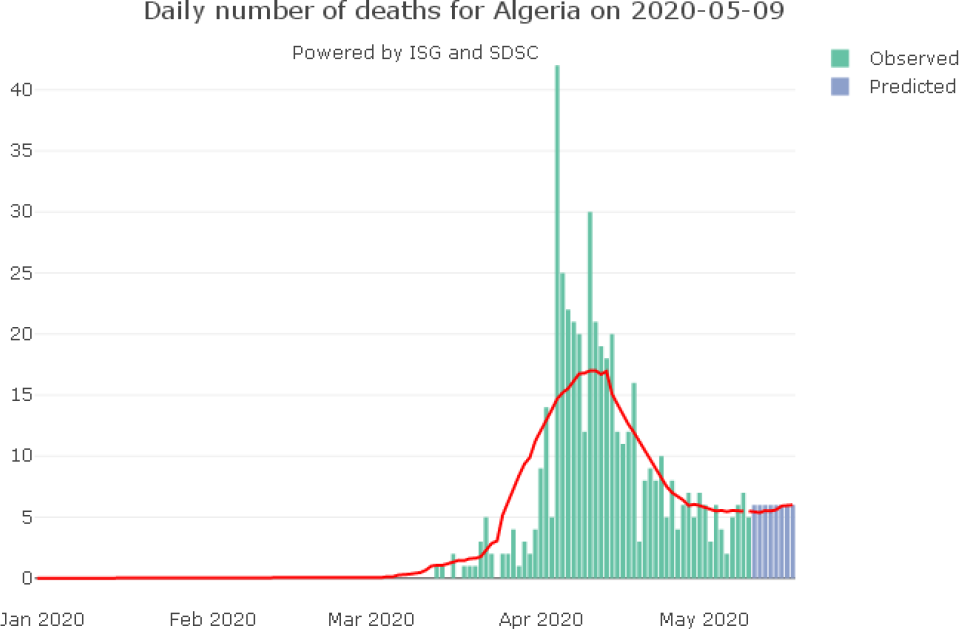
Daily number of deaths for Algeria on 09-05-2020 [17].

There is another study carried out on the propagation of Covid-19 in Algeria by applying the SIR model whose parameters were calculated based on the maximum likelihood estimation. This study done by [18] is very interesting because it deals with the same period as our work (25th February 2020 to April 24, 2020). Using their settings of the optimal SIR model which are: N (population size)= 3127, β (contact rate)= 0.18 and γ (removed rate)= 0.033; we get the following curve (figure 9) :

**Figure 9.**
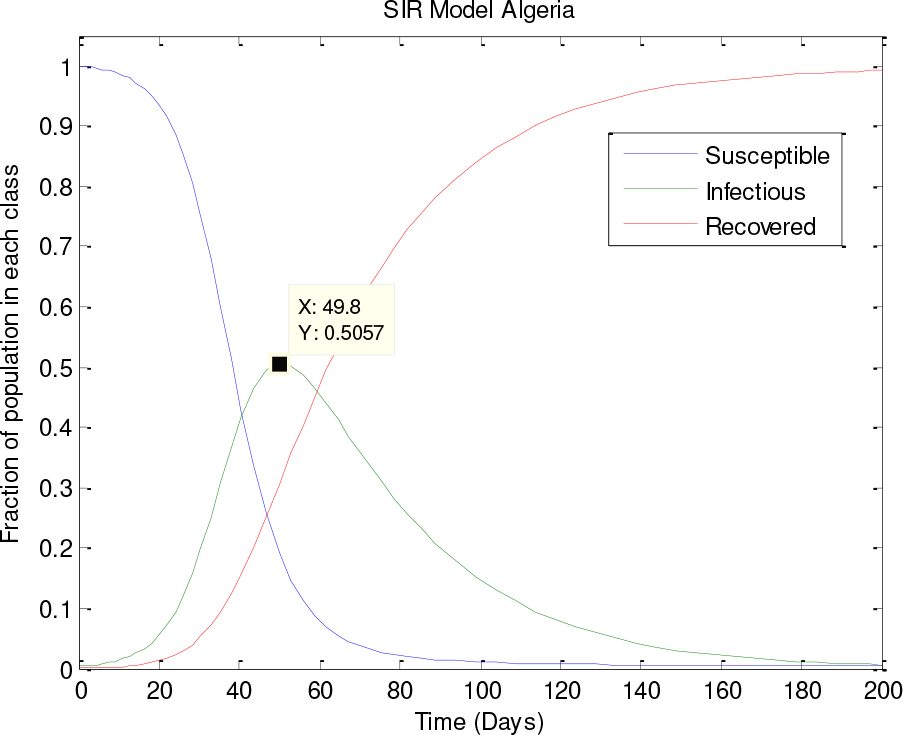
Temporal representation of SIR model variables in Algeria (peak time is 49.8 day≈50^th^ day).

From figure 9, it can be seen that the peak time corresponds to 50^th^ day (14th Aril) which gives the same result as the curve fitting (see section 3-1).

### 3.4 Drastic change in the situation for the later period

After a satisfactory evolution of the Covid-19 epidemic in Algeria in the period subject to this research, the containment measures enacted in Algeria were alleviated by a decision of the authorities taken on 24th April 2020. There was a bad reaction from the population and the situation had changed completely. At the time of completing this work (15th May), the extinction time of the epidemic has been extended and the infection curve has been shifted (figure 10).

**Figure 10.**
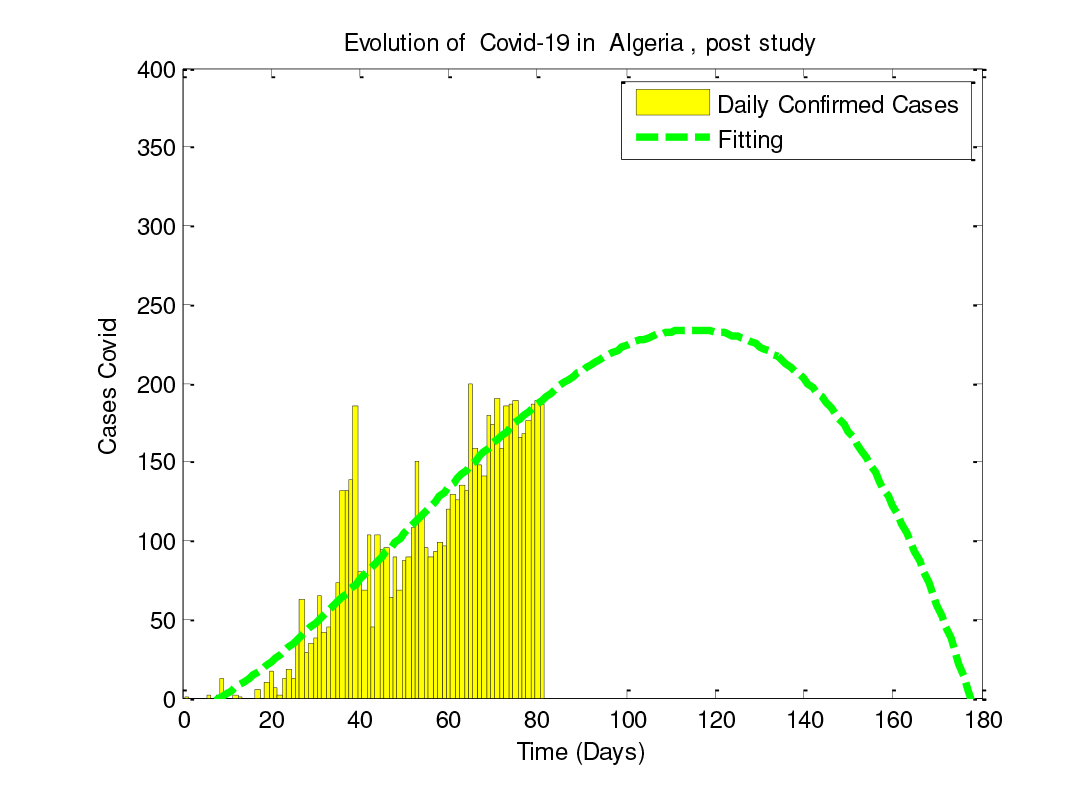
The time-Evolution by cubic polynomial fit of infected daily cases.

Subsequently, there has been a tightening of preventive measures by the authorities but it must take time for it to gives results.

## 4. CONCLUSIONS

An estimation of the time peak and total size of the coronavirus epidemic in Algeria is expected in this paper. The extent of the study period runs from 25th February 2020, to 24th April 2020. Our work has shown through several approaches (SIR model, curve fitting and statistical growth model) that the peak of this period was 10th April for some methods and 14th April for others. Although these two dates are very close, the date of 10th April is the most credible because it is given by the growth model based only on the real data without prediction.

The study of the spread of Covid-19 is still difficult and must have a margin of safety because it is considered as a “wicked problem” [19], given its uncertain nature depending on the social life of individuals as well as communities.

Moreover, for future work, we plan to apply other epidemiological models such as the SEIR model (Susceptible - Exposed - Infectious - Recovered) as well as the extension of the study period.

## Data Availability

/

